# Keeping weight off: Mindfulness-Based Stress Reduction alters amygdala functional connectivity during weight loss maintenance in a randomized control trial

**DOI:** 10.1101/2020.11.16.20226290

**Authors:** Serhiy Y. Chumachenko, Ryan Cali, Milagros C. Rosal, Jeroan Allison, Sharina Person, Douglas Ziedonis, Benjamin C. Nephew, Constance M. Moore, Nanyin Zhang, Jean A. King, Carl Fulwiler

**Author notes:** Corresponding author contact information., Address: University of Massachusetts Medical School, 55 Lake Avenue North, Worcester, MA, 01655, USA.

## Abstract

Obesity is associated with significant comorbidities and financial costs. While behavioral interventions produce clinically meaningful weight loss, weight loss maintenance is challenging. The objective was to improve understanding of the neural and psychological mechanisms modified by mindfulness that may predict clinical outcomes.

Individuals who intentionally recently lost weight were randomized to Mindfulness-Based Stress Reduction (MBSR) or a control healthy living course. Anthropometric and psychological factors were measured at baseline, 8 weeks and 6 months. Functional connectivity (FC) analysis was performed at baseline and 8 weeks to examine FC changes between regions of interest selected a priori, and independent components identified by independent component analysis. The association of pre-post FC changes with 6-month weight and psychometric outcomes was then analyzed.

Significant group x time interaction was found for FC between the amygdala and ventromedial prefrontal cortex, such that FC increased in the MBSR group and decreased in controls. Non-significant changes in weight were observed at 6 months, where the mindfulness group maintained their weight while the controls showed a weight increase of 3.4% in BMI. Change in FC at 8-weeks between ventromedial prefrontal cortex and several ROIs was associated with change in depression symptoms but not weight at 6 months. This pilot study provides preliminary evidence of neural mechanisms that may be involved in MBSR’s impact on weight loss maintenance that may be useful for designing future clinical trials and mechanistic studies.

## Introduction

As obesity rates in the United States continue to rise (Flegal, Kruszon-Moran, Carroll, Fryar, & Ogden, 2016; Fryar, Carroll, & Ogden, 2016), the number of obese adults is expected to rise in the US by 2030 by an additional 65 million (Wang, McPherson, Marsh, Gortmaker, & Brown, 2011). A risk factor for multiple serious disorders, including cancer (Renehan, Tyson, Egger, Heller, & Zwahlen, 2008), cardiovascular disease (Poirier et al., 2006), and metabolic syndrome (Dandona, Aljada, Chaudhuri, Mohanty, & Garg, 2005), obesity has established itself as one of the leading causes of morbidity and mortality both in the US and across the globe (Danaei et al., 2009; Forouzanfar et al., 2016). Furthermore, obesity is responsible for trillions of dollars in economic losses annually through medical costs, disability, and lack of productivity (Tremmel, Gerdtham, Nilsson, and Saha, 2017).

Weight loss can be an effective method to combat these obesity-related comorbidities (Knowler et al., 2002; Lavie, Milani, & Ventura, 2009; Wang et al., 2011), and a variety of effective weight loss strategies exist (Kushner, 2014). Maintenance of weight loss, however, remains more intervention-resistant (Merrill et al., 2008; Ory, Lee Smith, Mier, & Wernicke, 2010). Obese individuals typically regain one-third of the weight loss by the end of the first year following treatment (Wadden, Butryn, & Wilson, 2007; Rena R Wing, 2004), and by 3-5 years they have frequently returned to baseline weight (Franz et al., 2007). Thus, it is important to develop treatment strategies that address factors associated with weight regain.

Although findings are mixed (Elfhag & Rossner, 2005), there is compelling evidence from weight loss maintenance studies that psychological factors, particularly stress and depression, are significant predictors (Brantley et al., 2014; Elder et al., 2012; Trief, Cibula, Delahanty, & Weinstock, 2014; R R Wing et al., 2008). Failure to adequately address the impact of these factors on weight regain may explain the low success rates of existing interventions.

Mindfulness-based interventions have potential utility in weight loss and weight maintenance (Fulwiler, Brewer, Sinnott, & Loucks, 2015; Fulwiler et al., 2016). Mindfulness is defined as the awareness that arises from attending to one’s own internal and external state in the present moment in a non-judgmental way (Kabat-Zinn, 2003). Mindfulness meditation is a form of mental training focused on observing changing patterns of internal and external experience and reducing reactivity to aversive experience (Perlman, Salomons, Davidson, & Lutz, 2010; Zeidan, Grant, Brown, McHaffie, & Coghill, 2012) through enhanced self-regulation (Holzel et al., 2011; Loucks et al., 2015). The effectiveness of mindfulness-based interventions, in particular MBSR, for reducing aversive symptoms such as depression and anxiety is well-established (Goyal et al., 2014). Given that these symptoms increase the risk of unhealthy behaviors, MBSR may support maintenance of weight loss following successful initiation of health behavior change (Fulwiler et al., 2016).

Although studies of interventions for weight loss that incorporate mindfulness training have yielded mixed results, a recent systematic review and meta-analysis concluded that these interventions demonstrate moderate effectiveness (Carriere, Khoury, Gunak, & Knauper, 2018). However, the majority of these interventions involve varying combinations of meditation practices with training in mindful eating and elements of traditional weight management programs such as educational and/or behavioral skills training for improving diet and physical activity (Dalen et al., 2010; Miller, Kristeller, Headings, & Nagaraja, 2014; Miller, Kristeller, Headings, Nagaraja, & Miser, 2012; Timmerman & Brown, 2012). The contribution of mindfulness to the outcomes of these studies is unclear, leaving unanswered the value of the training in non-reactive, present-moment awareness for self-regulation (Fulwiler et al., 2015).

An important first step in studying interventions such as mindfulness is understanding the impact of the intervention on relevant neurobiological targets, and the relationship of this impact to clinical outcomes (Onken, Carroll, Shoham, Cuthbert, & Riddle, 2014). This type of mechanism-focused research can advance the study of behavioral interventions, and inform the design of clinical trials to test efficacy. For example, neuroimaging, including functional magnetic resonance imaging (fMRI), has proven useful in elucidating neurobiological mechanisms in neuropsychiatric disorders (Bruno et al., 2010; Charland-Verville, Habbal, Laureys, & Gosseries, 2012; Sambataro et al., 2010; Tshibanda et al., 2010), and, more importantly, the change in behavior and treatment response over the course of psychotherapy or other interventions (Crowther et al., 2015; McGrath et al., 2013; Walsh et al., 2017). Neuroimaging, including resting state (RS) fMRI, can also predict real-word dietary behaviors (Berkman & Falk, 2013; Giuliani, Merchant, Cosme, & Berkman, 2018)

Neurobiological changes associated with mindfulness training have recently been identified, pointing to possible biological mechanisms that could inform future clinical studies of effectiveness (Gotink, Meijboom, Vernooij, Smits, & Hunink, 2016). The best studied mindfulness intervention, Mindfulness-Based Stress Reduction (MBSR), is associated with functional connectivity (FC) changes in attention, sensory awareness, emotional regulation and dysregulation, and top-down control via the frontal lobe and the default mode network (Creswell et al., 2016; Kilpatrick et al., 2011; Roland et al., 2015; Taren et al., 2015). As such, fMRI could be an invaluable tool in identifying the impact of MBSR on neural circuit-level targets, thereby tailoring mindfulness interventions for individual effectiveness, tracking treatment progress, and optimizing overall outcomes at the population level.

Because fMRI allows for the observation of brain activation in real time via blood oxygenation level dependent (BOLD) imaging, it provides the opportunity to analyze the FC of various brain regions in an actively processing or RS brain (Biswal, Zerrin Yetkin, Haughton, & Hyde, 1995). This refers to the concept that regions of the brain that are functionally interconnected will co-activate in synchrony or predictable cadence, and thus temporal analysis of fMRI data can be used to determine which brain regions are functionally linked (van den Heuvel & Hulshoff Pol, 2010). Furthermore, higher-level FC analysis can establish functionally linked brain networks, or “independent components,” (ICs) of interconnected brain regions. This Independent Component Analysis (ICA) is a data-driven multivariate analysis technique that uses all of a given fMRI’s voxels in an image to simultaneously extract multiple ICs (Kiviniemi, Kantola, Jauhiainen, Hyvärinen, & Tervonen, 2003). Given its empiric, data-driven nature, ICA was the analysis method chosen in this study to extract specific functionally-derived networks of interest, where, in a standard approach similar to past studies (Fan et al., 2015) the connectivity of selected brain regions of interest (ROIs) into these networks could then be quantified.

We previously published our initial exploratory hypotheses and protocol for this randomized controlled trial (RCT) to investigate RS predictors of clinical outcomes following MBSR in the context of weight loss maintenance (Fulwiler et al., 2016; ClinicalTrials.gov (NCT02189187)). Our primary aim was to characterize RS changes in response to MBSR and the comparison condition. We hypothesized that MBSR would be associated with increased RS connectivity compared with the control condition. Based on previous fMRI research on MBSR, we selected several key brain regions of interest (ROIs) and ICs to focus our investigation (Garcia-Garcia et al., 2013; Hare, Camerer, & Rangel, 2009; McFadden, Cornier, Melanson, Bechtell, & Tregellas, 2013; Murdaugh, Cox, Cook, & Weller, 2012; Rothemund et al., 2007; Stice, Yokum, Bohon, Marti, & Smolen, 2010; Tregellas et al., 2011; Yokum, Ng, & Stice, 2011) (Fig.1). Our second aim was to investigate the association of RS change post-intervention with 6-month outcomes for psychological and anthropometric factors. We hypothesized that increased RS connectivity would be associated with improvement in depressive symptoms and inversely related to decreases in weight and waist circumference. In the current study, we present findings for our primary outcomes: 1) Change in RS fMRI signal following the 8-week MBSR intervention; 2) The association of RS fMRI changes with changes in weight (BMI) and depression symptoms at 6 months follow up. Exploratory analyses of other factors such as self-reported change in mindfulness, meditation practice time, and other psychological symptoms and health behaviors will be presented in future reports.

**Figure 1:**
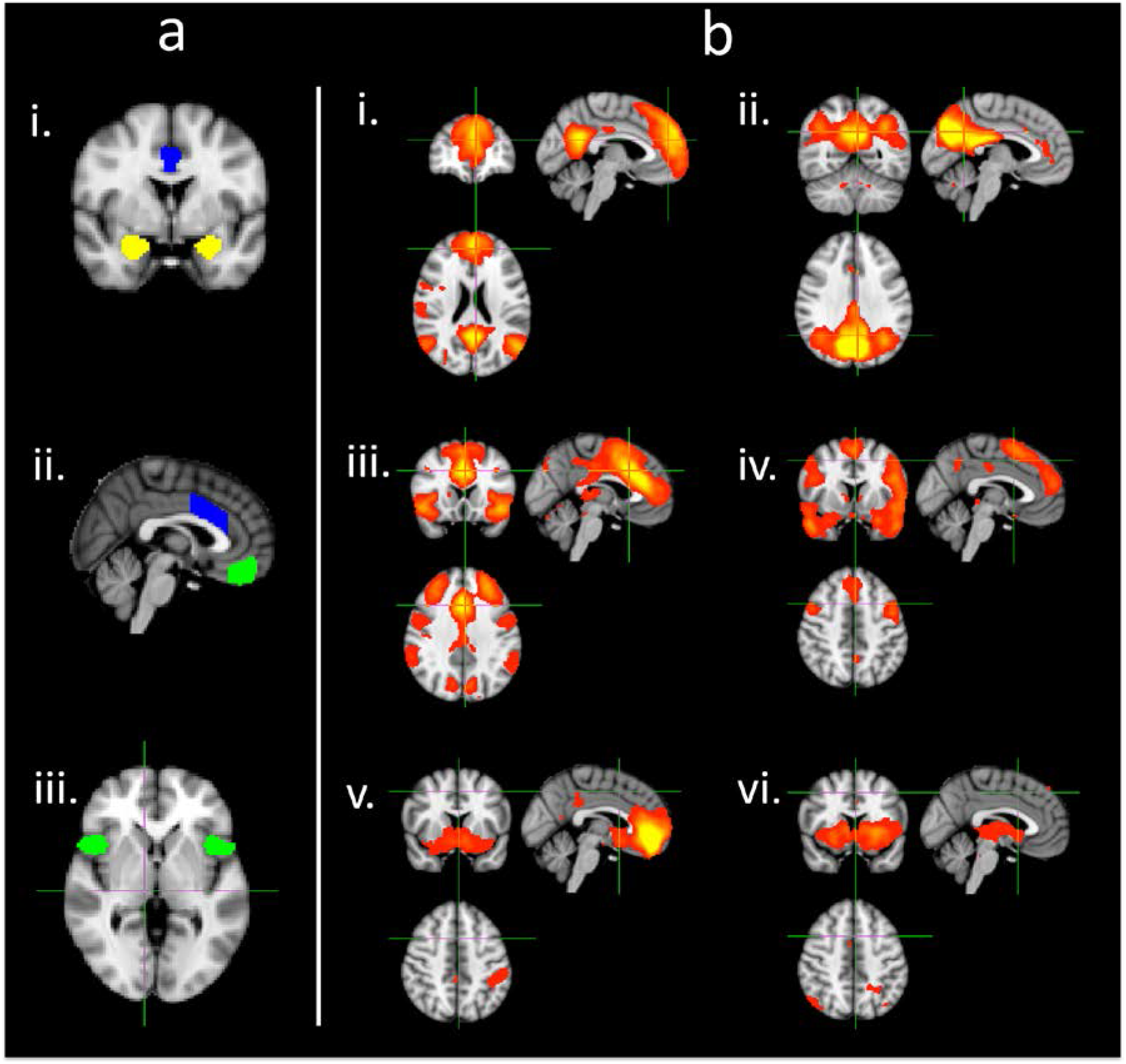
Selected ROIs (a) and independent components (b) a. ROls. Shown in each color: green (in image ii) -- medial OFC, blue (in image i and ii) -- dorsal ACC, yellow (in image i) -- amygdala, green (in image iii) -- anterior insula b. Independent components of interest as isolated as discrete ICs from MELODIC ICA. i: anterior DMN, ii: posterior DMN, iii: salience network, iv: dorsal DMN, v: vmPFC, vi: basal ganglia

## Methods

### Design and participant selection

The study was approved by the Institutional Review Board at the University Of Massachusetts Medical School. The study including primary hypotheses reported here was registered with ClinicalTrials.gov (NCT02189187). The overall study design and flow of subject selection are shown in Fig.2.

**Fig. 2.**
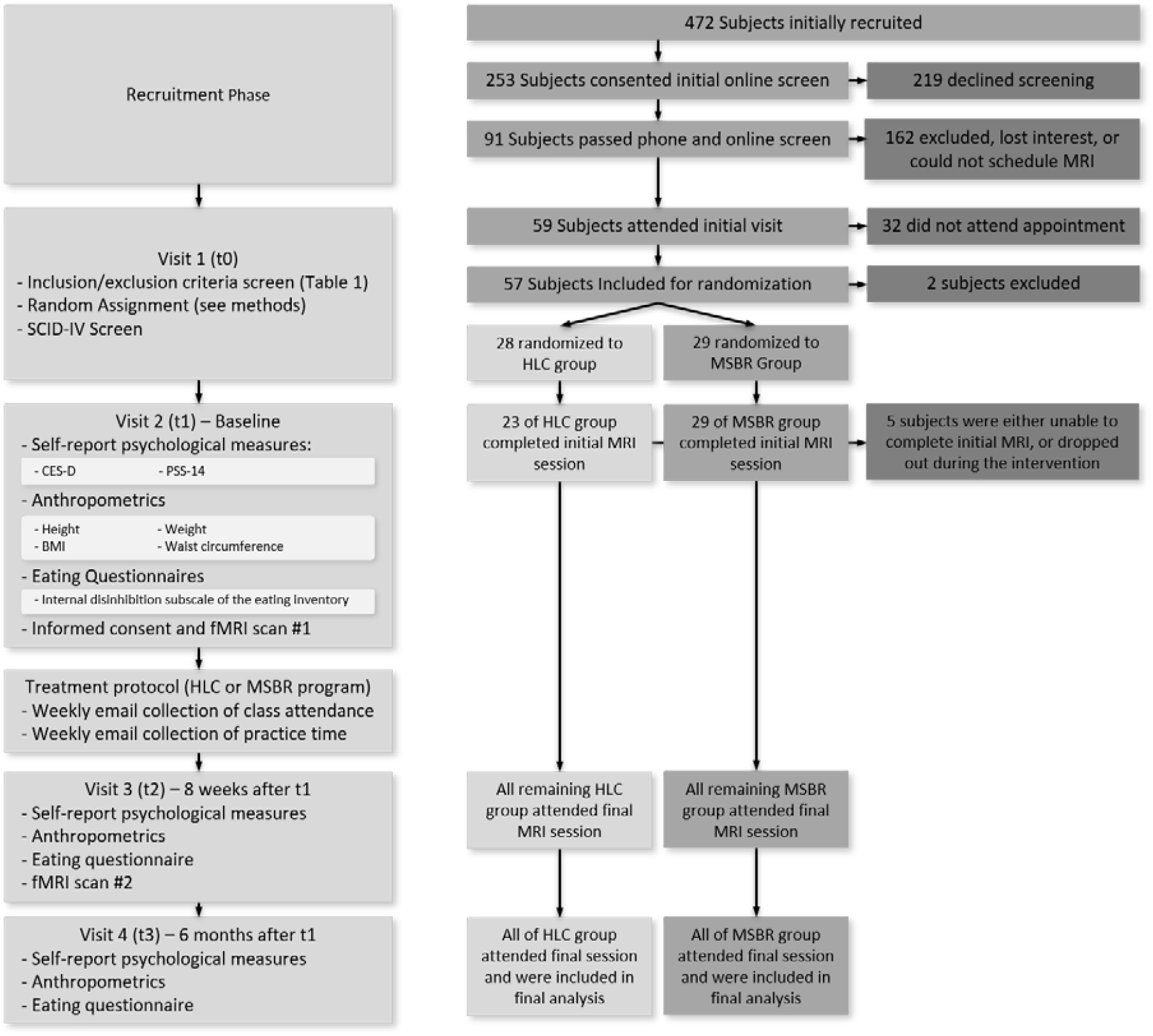
Study flow diagram. Left half of the figure shows the study phases, while the right half shows the flow of patients and numbers in each group at each of the corresponding study phase. This design is based upon our previously described model (Fulwiler et al., 2016)

Recruitment and screening procedures, including inclusion/exclusion criteria, and power calculation have been reported elsewhere (Fulwiler et al., 2016). Briefly, recruitment was conducted in the County of Worcester, MA, which has a population of approximately one million. We based study sample size on the primary outcome for functional connectivity change. The estimation indicated that 40 participants in each group were needed for 80% power.

Recruitment involved advertising (internet, flyers and social media), search queries through the UMass Memorial Medical Center electronic medical record and word of mouth. Potential subjects were directed to a web-based screening survey for initial eligibility screening and, if eligible, more detailed phone-based screening was scheduled. Patients of the UMass Memorial Medical Center authorized a Health Insurance Portability and Accountability Act (HIPAA) waiver.

472 subject were initially recruited to the study. A total of 253 individuals completed online screening for eligibility. 50 lost interest and did not complete the phone screen. Additional exclusions included: 115 individuals that did not meet the minimum 5% weight loss in the past year; 3 that were currently enrolled in a weight loss study, 8 that had a past weight loss surgery, 2 MRI exclusion, 7 who engaged in current meditation or yoga practice, 11 who had participated in an MBSR class previously, and 8 that had diabetes. Forty-two of those screened and found eligible declined to participate when told of the time commitment required for the 8-week classes, and 18 individuals could not be scheduled. Ninety-one participants passed all exclusion criteria and were invited to attend the initial visit. Of these, 59 attended the baseline visit and 57 were randomized.

Of the 57 subjects randomized, 5 were either unable to complete initial MRI, or dropped out during the 8-week intervention. 52 returned for the 8-week follow-up visit, and all 52 completed the 6-month follow-up visit (91% retention). 83% of participants completed 5 or more sessions, and 73% completed 6 or more.

### Randomization

Study participants, who were blind to the hypotheses, 57 were randomized to either the MBSR or the healthy living course (HLC) based on a permuted blocks randomization scheme (Fulwiler et al., 2016). Sample sizes consisted of 29 in the MBSR group and 28 in the HLC group. Randomization and investigators blinding was implemented using sealed envelopes and unique identification numbers by the study coordinator. In addition to the participants who were aware of which intervention they were receiving but not the study hypotheses, only the study coordinator among the research team was non-blinded to participant assignment until the study conclusion. MBSR teachers at the Center for Mindfulness were not aware of who in their classes were study participants. HLC teachers were aware that classes were being offered as part of the research study but were blinded to study hypotheses.

Interested subjects initially presented to the study site (visit 1) for initial informed consent for that visit, randomization, screening, and administration of a Structured Clinical Interview for the Diagnostic Statistical Manual IV (SCID-IV) interview. At visit 2, after written informed consent was obtained, subjects completed self-report psychological and behavioral measures, and anthropometric measures were obtained (see Fig.2), followed by a fMRI scan. Within 2 weeks of baseline data collection, participants started their assigned intervention (MBSR or HLC courses). Class attendance was recorded for participants in both conditions, and participants in the MBSR condition reported practice time. A second assessment was conducted within 2 weeks of completion of the interventions and included anthropometric and self-report measures and fMRI imaging. A third assessment was conducted 6 months after visit 2 and included self-reported and anthropometric measures only.

### Experimental vs Control Interventions

As described elsewhere (Fulwiler et al., 2016), the study included an experimental (MBSR) and an HLC control condition (Pbert et al., 2012), matched on course format (small group), length (8 weeks), number of sessions (eight 1.5 hour sessions plus a day-long retreat), small-group format, and process (didactic teaching and group discussion). The MSBR course was taught by certified teachers from the Umass Center for Mindfulness in eight weekly classes and one all-day retreat, with homework including formal meditation practices and informal practices during daily life. HLC classes were taught by trained health educators using a structured curriculum. HLC classes were audio recorded, and 10% of classes were reviewed for quality and fidelity to the curriculum.

### Assessments

Anthropometric and psychological measures (Fig.2) were administered at baseline, first follow up (within 2 weeks post-intervention) and second follow up (6 months following the first follow up). These measures are listed in Fig.2 and in the protocol paper. In addition, fMRIs were conducted at baseline and at the first follow up assessment. Due to claustrophobia and other contraindications, 5 subjects were not included in the MRI analysis as they did not undergo MRI, bringing the analysis to 51 subjects (MBSR N= 28, HLC N=23). All MRI data were acquired on a GE 3 Tesla scanner with a 12-channel coil. High resolution structural images were acquired using a using a magnetization-prepared rapid acquisition with gradient echo (MPRAGE) pulse sequence with the following parameters: TR/TE¼ 2.1 s/2.25 ms, slices¼ 128, matrix¼ 256 256, flip angle¼ 12, resolution¼ 1.0 1.0 1.33 mm. Gradient echo echo-planar images sensitive to BOLD contrast were acquired using the following parameters: TR/ TE¼ 2.0 s/30 ms, flip angle¼ 90, slices¼ 34, voxel size¼ 3.5mm isotropic. During the 6:04 minute RS fMRI scan participants were asked to remain awake with their eyes open FMRI and was carried out as described in our previous manuscript (Fulwiler et al., 2016). We used region of interest-based analysis to observe FC differences over time between regions and ICs of interest (see results section for further discussion).

### Data pre-processing

Prior to statistical analyses, preprocessing was performed on raw functional images using the FMRIB (functional MRI of the brain) Software Library (smith et al 2004; Woolrich et al 2009; Jenkinson et al 2012), (FSL) release 5.3 including: motion correction, slice-timing correction, non-brain removal and spatial smoothing (FWHM 5mm Gaussian kernel). ICA AROMA, an independent component analysis-based denoising tool, was then used to remove motion-related components and other components of no interest (e.g., respiration and artifacts) from the fMRI data (Pruim et al., 2015). No subjects were removed from the analysis due to excessive motion in the scanner. Denoised data were then temporally filtered using a Gaussian-weighted least-squares straight line fit with a high pass cutoff = 150 s. fMRI data were registered to MNI152 standard space is done in two steps. First, the fMRI data were aligned to the high-resolution structural image using 6 degree-of-freedom (DOF) affine transformation. Second, the structural scan was aligned with MNI152 standard brain using non-linear registration. Finally, transformation of the functional results into MNI space was done following concatenation of the two alignments into a single matrix. All spatially normalized fMRI data were re-sampled to 2mm^3^ resolution.

## Statistical Modeling

### Independent Component Analysis

FSL MELODIC [Multivariate Exploratory Linear Decomposition into ICs (Beckmann and Smith, 2004)] was used to perform an ICA of the fMRI data from all subjects and sessions (pre- and post-intervention). Pre-processed functional data was masked with MNI152 brain image to ensure exclusion of non-brain data, before inputting them into MELODIC. MELODIC decomposes the spatially normalized fMRI data from all subjects (e.g. concatenated into a single data matrix) into a set of ICs, with each IC being a distributed set of brain regions with a temporal trace that describes the evolution of that particular spatial pattern over time. The number of spatio-temporal patterns estimated by the group ICA was selected as the model order that gave resting state networks with the highest spatial cross correlation with the network spatial maps reported by Smith et al. (2009) which are available at https://www.fmrib.ox.ac.uk/datasets/brainmap+rsns/. The FMRIB Analysis Group at Oxford suggests that approximately 30 ICs is the “optimal” number to avoid overfitting and underfitting thus, 30 ICs were used in the analysis (Woolrich et al., 2009).

### Dual Regression

For group ICA, each IC is comprised of a spatial map and a corresponding time course and represents a spatio-temporal pattern of activity that is common to the entire set of participants. To estimate the spatial patterns and time courses for each subject (which capture the between-subject variability), dual regression (Nickerson et al., 2017) was applied as follows. Before conducting dual regression, the melodic maps were thresholded to ± Z score of 2.3 (cluster significance P < .05). Each IC was then normalized to maximum value 1 by dividing each component by its maximum Z score to account for differences in the scale of the spatial maps. In the first stage of dual regression, the group ICA maps were regressed onto each subject’s fMRI data to identify the time courses of each network within each participant. In the second stage of the dual regression, these time courses are normalized to unit variance and are then regressed against the participant’s fMRI data to estimate the unique set of network maps in each participant.

### Group analysis

As our objective to investigate the connectivity changes of *a priori* ROIs (medial OFC, dorsal ACC, amygdala, anterior insula) with *a priori* networks (anterior DMN, posterior DMN, salience network, dorsal DMN, vmPFC and basal ganglia) with intervention, we extracted connectivity values (average regression weights) of each ROI from the subject-specific spatial maps output from the second stage of dual regression for each subject. A group x time x hemisphere repeated measures ANOVA was then carried out for each of the ROI-IC pairs. All analyses were performed in SPSS v24.

## Results

### Baseline group comparisons

Table 1 describes baseline characteristics of participants in the MBSR and the HLC study conditions. No significant differences were found in baseline demographic, anthropometric, psychological or behavioral measures (Table 1).

**Table 1:** Sample characteristics of the 57 included study participants. Abbreviations: MBSR: mindfulness-based stress reduction, HLC: healthy living course, CESD: Center for Epidemiologic Studies Depression scale, PSS-14: Perceived Stress Scale 14, BMI: body-mass index.

|  | MBSR Group | HLC Group |
| --- | --- | --- |
| *Demographic Characteristics* |  |  |
| *N* | 29 | 28 |
| Age | 44.5 SD= 9.70 | 44.5 SD= 10.65 |
| Sex (% Female) | 76% SD= .43 | 87% SD= .34 |
| Race (% White) | 79% | 87% |
| Mean years of education | 17.2 SD= 2.23 | 17.0 SD= 2.32 |
| Handedness (% Right-handed) | 90% SD= .30 | 70% SD= .47 |
| *Baseline Questionnaire Characteristics* |  |  |
| CESD | 5.79 SD= 3.85 | 7.93 SD= 5.45 |
| PSS-14 | 16.52 SD= 3.48 | 20.14 SD= 3.24 |
| Internal disinhibition subscale of the Eating Inventory | 2.48 SD= 1.83 | 3.15 SD= 2.10 |
| *Baseline Anthropometric Characteristics* |  |  |
| Weight (lbs) | 177.1 SD= 36.39 | 178.8 SD= 39.29 |
| Waist Circumference (cm) | 93.1 SD= 11.22 | 93.1 SD= 11.95 |
| BMI | 28.4 SD= 4.50 | 29.4 SD= 5.90 |

### Change in resting state connectivity following the 8-week interventions

The connectivity between each of the 4 ROIs and each of the five ICs of interest (namely the estimated marginal means values) were analyzed individually for a group x time interaction using repeated measures ANOVA. A third category of hemisphere was added to the interaction to assess lateralization. A group x time interaction was found between the amygdala and vmPFC (Fig.3) (p=0.046 CI = 95%, .064, 7.61), with no interaction with hemisphere (p > 0.6), showing that FC was strengthened in the MBSR and reduced in the HLC control group over the 8-week course. No significant main effect of group or hemi x group was observed. Post-hoc analyses revealed that this interaction was driven by a significant group difference at 8 weeks (p=0.010) rather than the non-significant baseline differences (p>0.6). No other significant group x time interaction was found for any other ROI x component of interest, with or without an effect of hemisphere. Additionally, there were no additional significant effects found after removal of interaction effects.

**Fig. 3.**
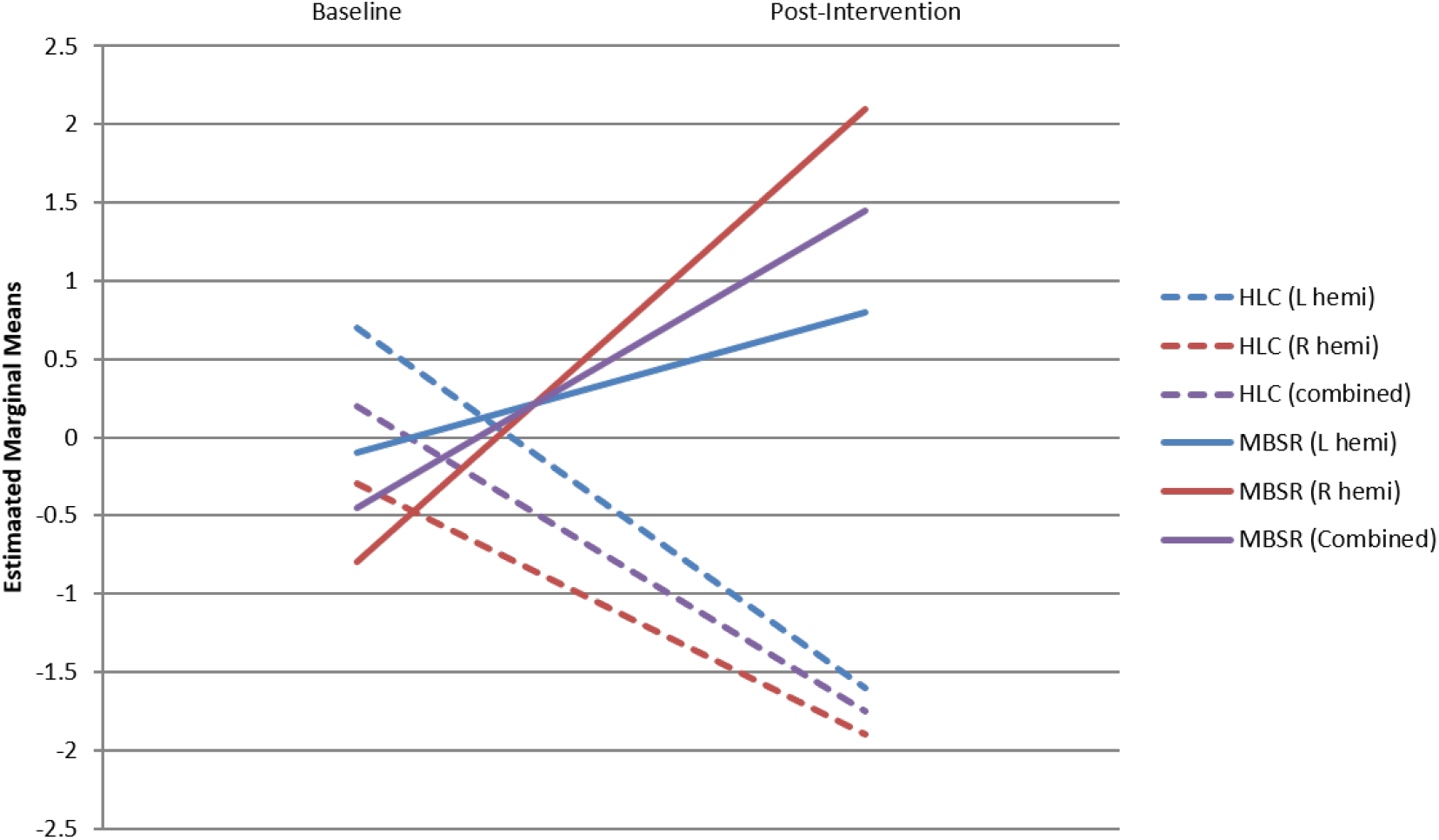
Change over time in functional connectivity between the amygdala and the ventromedial prefrontal cortex, in the left (blue lines), right (red lines) and combined (purple lines) hemispheres (hemi) of both the Healthy Living Course (HLC) group (dashed lines) and Mindfulness-Based Stress Reduction course (MBSR) group (solid lines). Higher estimated marginal mean values denote higher functional connectivity.

### Association of connectivity and 6-month anthropometric and psychometric outcomes

There were no statistically significant changes in the measured anthropometric metrics (weight, BMI, waist circumference) and psychological variables (depression, stress, internal disinhibition) from baseline to 6-month follow up. However, post-hoc analysis revealed that the mean weight of MBSR participants remained about the same (losing 0.7 lbs, or 0.4% of body weight and 0.1kg/m^2^, or 0.5% of BMI, and 3.2cm, or 3.5% of total waist circumference), while increasing in the HLC group (gaining 5.9 lbs or 3.2%, BMI by 1.0kg/m^2^ or 3.4%, and waist circumference by 1.2cm or 1.2%). No statistically significant differences between the groups were observed in psychological or eating measures.

Our second primary aim was to investigate the association of FC change with changes in psychological and anthropometric outcomes (Fulwiler et al., 2016). A paired t-test of the association between connectivity change at 8 weeks and weight and depression at 6 months did not reveal between-group differences. However, using a single all-subject analysis combining both groups and standard linear regression, a significant correlation was found for decrease in FC at 8-weeks (between vmPFC and a cluster of structural regions that contained the precuneus, inferior frontal gyrus, and parietal operculum), and increased CESD score change (in either direction) at 6 months (p = 0.02). Other significant correlations were found between increased 8-week FC change in basal ganglia and the cluster consisting of hippocampus, nucleus accumbens, and OFC and increased 6-month CESD change (p = 0.032); and increased 8-week FC change in the salience network and occipital and inferior temporal gyrus and increased CESD change (p = 0.02). Conversely, lower connectivity between ventromedial prefrontal cortex and the cluster containing precuneus, inferior frontal gyrus, and parietal operculum was associated with increased CESD score change (p = 0.02). No associations were found between FC changes and 6-month anthropometric changes.

## Discussion

The goal of the present study was to identify neural processes modified by MBSR that may be associated with weight loss maintenance and related emotional symptoms. We also examined changes in RS following MBSR vs. a rigorous control condition and, in an exploratory analysis, examined the association of these changes with depression symptoms and weight stability at 6-month follow-up. We confirmed our hypothesis that change in functional connectivity would be greater in the MBSR group compared to the comparison group. Specifically, this significant effect for FC changes was for amygdala-vmPFC connectivity, in agreement with previous studies. Secondly, our exploratory analysis of the association between FC change at 8 weeks and clinical outcomes found that FC change is associated with change in depression symptoms at 6-month follow-up.

### Neural targets of mindfulness and relevance to weight loss studies

Our findings are consistent with studies in other populations which show that mindfulness training leads to changes in medial prefrontal-amygdala connectivity critically implicated in the regulation of emotion. In one study, a mindfulness strategy for regulating emotional response was associated with regulation of amygdala activation by medial prefrontal cortex, in contrast to emotion suppression involved connectivity with dorsolateral prefrontal cortex (Murakami et al., 2015). In other studies, MBSR training led to decreased amygdala reactivity accompanied by increased amygdala-vmPFC functional coupling (Kral et al., 2018) and improvement in anxiety symptoms that were correlated with increased amygdala-vmPFC coupling (Hölzel et al., 2013). Considering these reports of increased functional coupling, our finding of increased RS connectivity following MBSR strengthens the evidence for the importance and specificity of the amygdala-vmPFC connection as a marker of MBSR treatment progress, efficacy, and outcomes.

The impact of mindfulness on these pathways may be particularly relevant to understanding how mindfulness-based interventions may be helpful for sustaining the behavioral changes required for weight loss maintenance, especially in light of evidence that stress and negative emotion disrupt the neural network supporting response inhibition (Patterson et al., 2016). Training to improve attentional focus through mindfulness meditation includes attending to thoughts and emotions while avoiding habitual reactivity to them. In addition to its role in emotion generation, the amygdala plays an important role in the salience network, placing it at the intersection of goal-directed behavior, “wanting”, and emotional triggers of habitual responses. The vmPFC has been implicated in self-control and reward processing through extensive connections with the amygdala and other subcortical structures (Bari & Robbins, 2013; Hare et al., 2009; Ochsner, Silvers, & Buhle, 2012).

Mindfulness training has been shown to modulate value signals in the vmPFC and reduce activation in response to reward (Kirk, Brown, & Downar, 2015; Kirk, Gu, Harvey, Fonagy, & Montague, 2014). The concept that mindfulness strengthens top-down executive control is not novel, and has been extensively discussed in the literature (Chiesa, Serretti, & Jakobsen, 2013; Creswell, Way, Eisenberger, & Lieberman, 2007; Hölzel et al., 2013; Lutz, Slagter, Dunne, & Davidson, 2008; Teper, Segal, & Inzlicht, 2013). However, to our knowledge this is the first report of connectivity change following mindfulness training among individuals who have recently achieved weight loss. Top-down executive control of subcortical structures including the amygdala and striatum by PFC mediates goal-directed behavior and is disrupted by stress or negative emotion, allowing the salience network to drive behavior, promoting habitual emotional responses (Arnsten, 2009). Mindfulness meditation is believed to enhance emotion regulation at least in part through its effect on executive control to maintain guidance of behavior in line with personal goals over impulsive reactions to unpleasant emotions (Teper et al., 2013).

For vulnerable individuals, the habitual response to negative or stressful emotions may be eating palatable food which provides immediate, albeit short-term, relief (Dallman, 2010). Recent evidence provides support for the amygdala and PFC as key links between emotion dysregulation and eating behavior (Rothemund et al., 2007; Stice et al., 2010; Yokum et al., 2011). Amygdala activity increases in response to images of high-calorie foods compared to non-food images (Murdaugh et al., 2012), in line with its role in the reward pathway. Furthermore, activation of both the amygdala and the vmPFC is increased after the consumption high-fat versus lean foods (Ng, Stice, Yokum, & Bohon, 2011). The PFC is implicated in self-control, and decreased activity of the medial PFC predicts weight gain in obese individuals (Kishinevsky et al., 2012). Thus, for an experimental medicine approach to refining mindfulness interventions for emotion regulation and behavior change research (Onken et al., 2014), neuroimaging markers such as connectivity between amygdala and medial PFC may prove useful for optimizing the intervention and identifying subpopulations most likely to respond (see Gabrieli et al. (Gabrieli, Ghosh, & Whitfield-Gabrieli, 2015) for a review of the neuromarker approach). As obesity is the cumulative result of a range of specific eating behavior traits (Carnell, Gibson, Benson, Ochner, & Geliebter, 2012), understanding neural targets specific to different subgroups will be essential. For example, we have reported preliminary evidence that MBSR leads to reduced emotional eating especially for those with high emotional eating scores at baseline(Levoy, Lazaridou, Brewer, & Fulwiler, 2017). There may be specific neural targets for emotional eaters vs. other subtypes of overeaters.

### Association of connectivity changes with clinical outcomes

Our second primary aim was to examine whether pre-post change in FC would predict 6-month change in weight and depression symptoms. A correlation with weight was not found. On the other hand, we report for the first time that FC change is associated with change in depression symptoms at 6-month follow-up. This finding was not specific to MBSR as our sample size was too small to detect differences between groups. Given this sample size issue, despite non-significant changes over time in our collected anthropometric and psychologic measures, this lack of statistical significance does not necessarily preclude real group differences in these anthropometric and psychologic measures. The finding included several ROI’s but did not include amygdala-vmPFC connectivity.

Depression symptoms were chosen as a clinical measure because they have been linked to unsuccessful weight loss and weight loss maintenance outcomes, although findings have been mixed (Brantley et al., 2014; Konttinen, Silventoinen, Sarlio-Lähteenkorva, Männistö, & Haukkala, 2010; Ouwens, van Strien, & van Leeuwe, 2009; Teixeira, Going, Sardinha, & Lohman, 2005), and are a marker of the effect of MBSR on emotion regulation. However, our recent systematic review of emotion regulation measures in mindfulness studies found that the CESD was not as reliable as measure of depression symptoms compared to clinician-rated scales (Kimmel, et al.; manuscript in preparation).

That amygdala-vmPFC connectivity change was not associated with either depression symptoms or weight metrics at 6 months indicates that this marker may not have utility for mindfulness interventions in the general weight loss population. However, future studies may examine its association with clinical outcomes in specific subpopulations. In contrast, vmPFC connectivity with other areas such as a cluster that included the IFG was correlated with depression symptoms. The IFG plays a critical role in attentional control and various forms of response inhibition, including emotional inhibition (Aron, Robbins, & Poldrack, 2004), and disruption of IFG FC by negative emotion is believed to be responsible for the adverse effect of negative emotion on response inhibition (Patterson et al., 2016). Further work is warranted on the roles of both amygdala and vmPFC connectivity in behavioral mechanisms of weight loss.

### Study strengths and limitations

This study had a number of strengths. The two groups were well-matched in terms of baseline demographic, psychometric and anthropometric characteristics, with no statistically significant baseline differences. The comparison condition was well-matched to the experimental condition on course length, style, and number of sessions, employing both a small-group format that included didactics and group discussion. In addition, we employed a 6-month follow-up for testing the effect of interventions on clinical outcomes.

A significant limitation is that recruitment goals were not achieved resulting in the trial being underpowered. Our primary outcomes were powered on a sample size of 80 participants, but only 52 participants completed the study. Thus, while the sample size allowed us to detect a significant group difference in functional connectivity change in pre-selected regions of interest in the MBSR group as predicted, between group changes in clinical outcomes were not significant and we were unable to confirm the hypothesis that functional connectivity changes would predict clinical outcomes in the MBSR group. This limitation is not uncommon in trials involving mindfulness-based interventions, in part because the intervention demands a large investment by participants. After expressing initial enthusiasm for the study, many potential recruits declined to participate because of the time requirement for the interventions.

Another limitation of the study is the largely white and female demographics of the sample, especially in light of evidence that race, sex, and socioeconomic status may strongly affect weight loss outcomes (Ball & Crawford, 2005; Davis, Clark, Carrese, Gary, & Cooper, 2005; Hartmann-Boyce, Jebb, Fletcher, & Aveyard, 2015). In addition, although baseline differences between groups were not significant, the magnitude of differences in some variables may have been important and should be adjusted for in future studies with a larger sample. Our initial hypotheses were exploratory, so FC between several regions of interest and independent components were tested. Thus, a limitation of this study is the use of multiple testing which may inflate false positives. Furthermore, this study did not address the continued longitudinal follow-up of FC throughout weight loss maintenance, and whether long-term success in weight loss maintenance is correlated to connectivity markers. In addition, the study did not establish a quantitative approach to using FC as a biomarker. A more in-depth approach correlating clinical measures to FC would be needed to establish amygdala-PFC connectivity as a measure with clinical utility. Finally, without data on participants’ practice of mindfulness between sessions, we do not know the actual “dose” of mindfulness received.

## Conclusions

The present study provides evidence that mindfulness training in people who have lost weight and intend to keep it off leads to increased RS FC in a neural circuit involved in emotion regulation. These findings are consistent with previous research suggesting that short-term training in mindfulness meditation alters the neural circuitry of emotion regulation, specifically connectivity between the amygdala and vmPFC. While connectivity change in this circuit was not associated with later clinical outcomes, preliminary evidence suggests an association between other vmPFC connectivity changes and depression symptoms which may be relevant to weight loss maintenance.

## Data Availability

Data is available through the Open Science Framework.

https://osf.io/q3rch/

## Acknowledgements and Funding

The authors thank Drs. Shaokuan Zheng, Wei Huang, Guillaume Poirier, and Poornima Kumar for technical assistance; Dr. Asimina Lazaridou, Marcela Hayes, Emily Levoy, Julia Siegel, Garen Koutoujian, Mark Fusunyan, Dr. Gertrude Manchester, and Dr. Nivedita Gour for their helpful contributions to the study. We thank Drs. Lori Pbert and Sarah Reiff-Hekking, developers of the Healthy Living Course, for sharing their expertise with the intervention. We thank the Conquering Disease Program funded by UMCCTS grant UL1TR000161 for help with recruiting. We also extend our sincere gratitude to the staff and teachers of the Center for Mindfulness for their assistance. The project was conducted at the UMMS Advanced MR Imaging Center.

The National Center for Complementary and Integrative Health (NCCIH) provided funding for this trial (grants R34AT006963 and R34 AT006963-01A1 to CF and JAK). The design of this trial was reviewed and approved by NCCIH’s Office of Clinical and Regulatory Affairs. Support was also provided by an award from the UMass Medical School (UMMS) Department of Radiology. The content is solely the responsibility of the authors and does not necessarily represent the official views of the National Institutes of Health or the UMMS Department of Radiology Advanced MR Imaging Center.

---

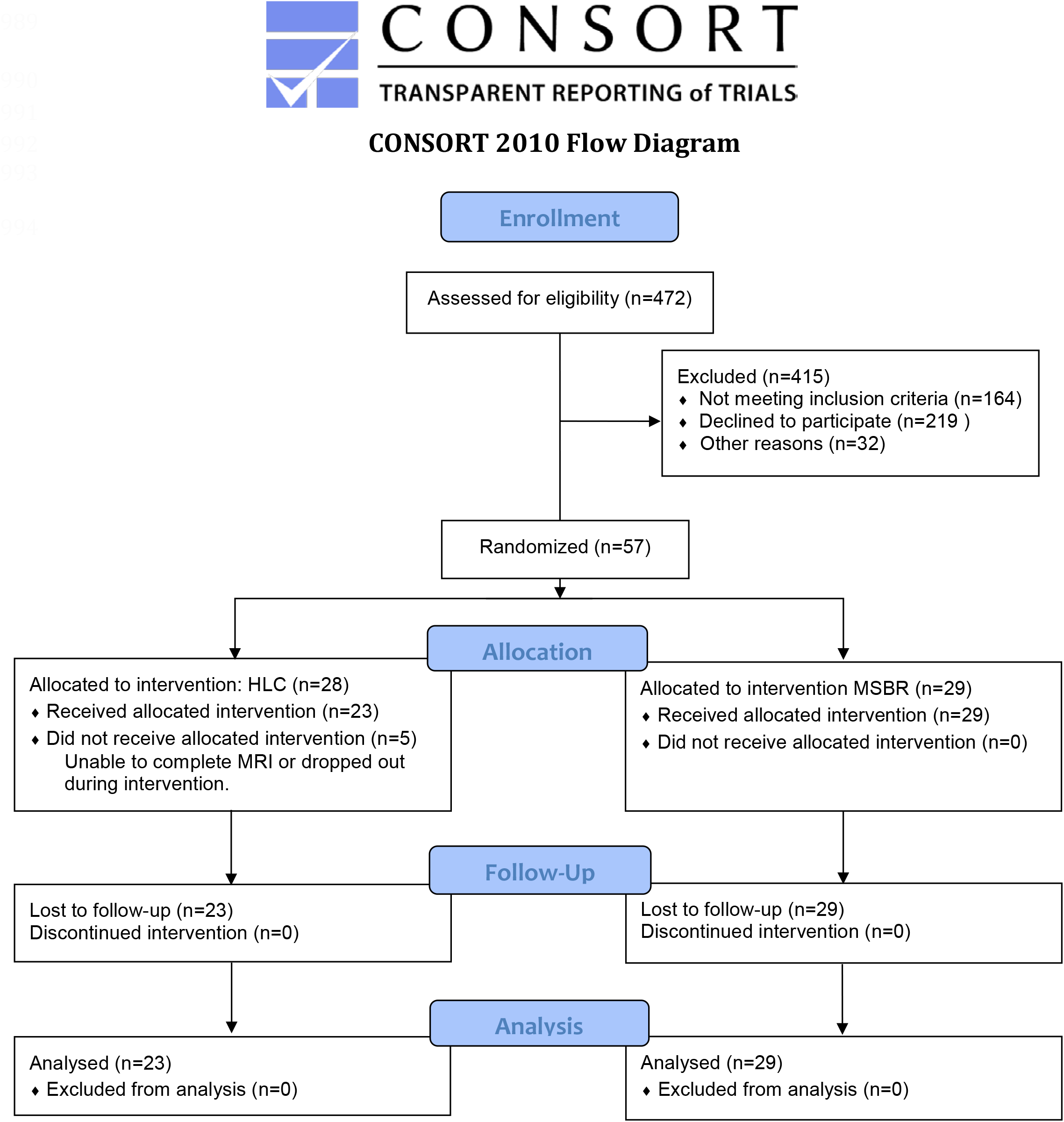

## Notes

### Competing Interest Statement

The authors have declared no competing interest.

### Clinical Trial

NCT02189187

### Clinical Protocols

https://bmjopen.bmj.com/content/6/11/e012573

### Author Declarations

The study was approved by the Institutional Review Board at the University of Massachusetts Medical School. The study including primary hypotheses reported here was registered with ClinicalTrials.gov (NCT02189187).

